# Monkeypox virus-neutralising antibodies detected against Clade Ib and Clade IIb in healthy individuals following MVA-BN vaccination

**DOI:** 10.1101/2025.09.17.25335983

**Authors:** Victoria H Sheridan, Craig W Duffy, Jake Dunning, Lance Turtle, Julian A Hiscox, Krishanthi S. Subramaniam, the ISARIC4C Investigators

**Author notes:** **Corresponding Author:** Dr Krishanthi Subramaniam, Department of Clinical Infection, Microbiology and Immunology, Institute of Infection, Veterinary and Ecological Sciences, University of Liverpool, Liverpool UK L69 7BE.

## Abstract

Recently, a novel subclade of mpox virus (MPXV), clade Ib, has emerged in Eastern Democratic Republic of the Congo (DRC). Modified vaccinia Ankara—Bavarian Nordic (MVA-BN) is a third-generation smallpox vaccine that is authorised and in use as a vaccine against mpox. MVA-BN provides approximately 78% protection against disease to Clade IIb MPXV, but there are no data for Clade Ib. We used a plaque reduction neutralisation assay (PRNT) with MPXV clinical isolates from Clade Ib and IIb to investigate immunity in vaccinated health care workers. Here we show that a two-dose regimen of MVA-BN can induce MPXV-neutralising antibodies against both clades, with higher responses to Clade IIb than Clade Ib.

**KEY POINTS:**

1. Vaccination with MVA-BN generates antibodies that can neutralise MPXV Clade Ib and IIb.
2. Despite the small sample size paired analysis across the vaccine recipients demonstrate higher neutralisation to Clade IIb than Ib.

## INTRODUCTION

Mpox is a zoonotic viral disease caused by mpox virus (MPXV), divided into Clades I (originally Central African) and II, with Clade II further subdivided into subclades IIa and IIb ^1,2^. In 2023 a new subclade of Clade I, termed Clade Ib, emerged in the Eastern Democratic Republic of the Congo (DRC). Since the first human case discovered in August 1970 in DRC, mpox has been reported in 11 African countries, with a small number recorded outside of Africa. However, in 2022 a global outbreak occurred in nonendemic areas caused by the Clade IIb strain^1^. More recently, infections associated with MPXV Clade Ib have been spreading and became a Public Health Emergency of International Concern (PHEIC) in August 2024. Clade Ib is predominantly transmitted through close household contact, particularly affecting children and has been linked to a higher number of deaths^12^. This variant also has higher transmission rates in comparison to Clade IIb, possibly due to its APOBEC3 mutation, causing a rapid evolution of the virus^12^. The WHO recommends those at high-risk of contracting mpox, especially during an outbreak receive vaccination^2^ with the modified Vaccinia Ankara-Bavarian Nordic (MVA-BN) smallpox vaccine, which is a live attenuated vaccine used worldwide, including in the UK, Europe, and the USA^1^.

Evidence demonstrates that vaccination with MVA-BN can generate low levels of neutralising antibodies against Clade IIb and Clade Ia^3,4^. In the UK, a single dose of MVA-BN provides short-term protection of 78% against mpox predominantly in the MSM (men who have sex with men) community^13.^ However, whether vaccination can also induce neutralising antibodies to Clade Ib has not been addressed. Using a small cohort (n=25) of healthy adults vaccinated with MVA-BN due to occupational exposure to mpox this study measured the neutralising antibody against Clades Ib and IIb using a plaque neutralisation assay (PRNT). Our results provide the first evidence that MVA-BN vaccination can elicit antibodies that can neutralise Clade Ib, albeit at low levels, and provides comparative data against Clade IIb for which the vaccine is known to be effective^13^. This finding has public health significance as it shows that vaccine recipients mount antibody responses that neutralise currently circulating MPXV clades. This study is important to assist policymakers with decisions regarding further vaccinations and whether a third vaccine dose is required.

## METHODS

### Study design and participants

Blood samples were obtained from twenty-five healthcare workers with occupational exposure to mpox who received two doses of the Modified Vaccinia Ankara (MVA) Bavarian Nordic (MVA-BN) vaccine via subcutaneous or intradermal routes (Supplemental Table 1). Vaccine recipients were recruited into the Acute Virus Immunity Study (AVIS; REC 16/NW/0170) at the University of Liverpool. All participants provided written informed consent before sample collection.

### Viruses and Cells

The Clade IIb mpox virus, 2022, Slovenia ex Gran Canaria was obtained from the European Virus Archive Global. MPXV Clade Ib, 2024, London ex East Africa (October 2024) was isolated from a clinical sample collected under the ISARIC Clinical Characterisation Protocol-UK (approved in England by Oxford Research Ethics Committee, ref 13/SC/0149). We passaged virus stocks in Vero E6 cells (African green monkey kidney, C1008 Public Health England PHE – UK Health Security Agency (UKHSA)) maintained in Dulbecco’s Modified Eagle Medium (DMEM) with 2% heat-inactivated fetal bovine serum and 0.05mg/ml gentamicin and harvested 72 h post inoculation.

### Virus Sequencing

DNA was extracted from MPXV stocks using Zymo DNA/RNA viral extraction kit. The NextGen PCR MPXV Sequencing Library Prep Kit (MBS) was used in conjunction with Oxford Nanopore Rapid Sequencing DNA kit V14 - barcoding (SQK-RBK114.24) to amplify viral DNA^15^. Mpox PCR amplicons were sequenced on an Oxford Nanopore P2 R10.4 flow cell following manufacturer’s instructions.

### Plaque Reduction Neutralisation Titres (PRNT)

PRNTs were performed using Vero E6 cells. Sera were diluted 1:4 in DMEM (2% FBS; 0.05 mg/mL gentamicin) followed by serial twofold dilutions. MPXV at 700 PFU/mL was added to an equal volume of diluted sera and incubated at 37°C for 1 h. In some experiments, 10% guinea pig serum (a source of complement), was added to the starting serum dilution (final concentration 5% when mixed with MPXV). The virus-serum dilution was inoculated onto Vero E6 cells in duplicate. Cells were incubated for 72 h at 37°C and 5% CO2 before being fixed with 10% formalin and stained with crystal violet solution. The 50% plaque reduction titre (PRNT_50_) was determined via probit regression analysis using a script adapted from Bewley et al. 2021^7^.

### Statistical Analysis

Statistical analysis was done using GraphPad Prism version 10 (GraphPad Software). The non-parametric Mann-Whitney U-test and Wilcoxon matched-pairs signed-rank tests were used to compare differences across the samples. P values less than 0.05 were considered significant.

## RESULTS

The identity of viral stocks was confirmed using Oxford Nanopore sequencing and alignment against previously published data (Supplementary Figure 1). The level of neutralising antibodies was measured in twenty-five adults with a median age of 39 years (interquartile range, IQR = 30 – 45). Sixteen of the 25 study subjects were female, and the ethnicity of the cohort included White (n=20), Asian (n=3) and Latin (n=2). Three individuals reported the following comorbidities: multiple sclerosis, psoriasis and asthma.

The importance of complement in relation to neutralisation levels has been reported for MPXV^6^ in addition to other viruses^8,9^. To assess the contribution of complement in our cohort, serum samples were exposed to different conditions (heat-inactivation (HI), HI supplemented with guinea pig serum as a complement source, and non-HI). Here we found, as previously reported^6^, complement is required for neutralisation of MPXV *in vitro* (Figure 1). Additionally, no significant difference in MPXV neutralisation was detected (p=0.0625, Wilcoxon signed-rank test) between HI serum in the presence of a complement source and non-HI. Based on these data we used non-HI serum for the remainder of the experiments.

**Figure 1.**
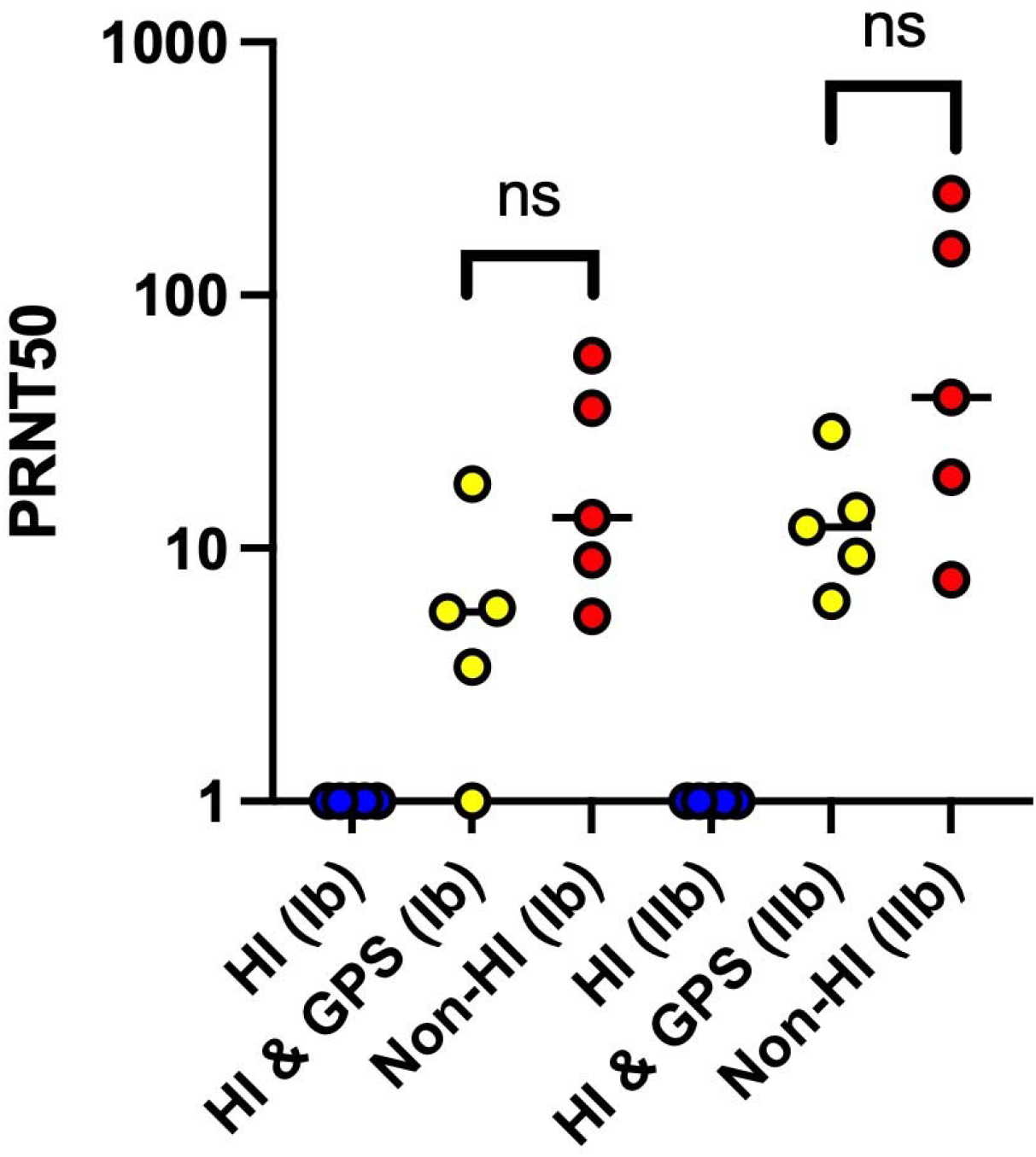
Log PRNT titres for participants vaccinated with 2 doses of MVA BN vaccine to assess neutralising antibody responses to MPXV Clade Ib and Clade IIb under different conditions to assess inclusion of complement. Each datapoint represents the geometric mean titre of 2 experimental replicates. Black line represents median. MPXV, monkeypox virus; PRNT, plaque reduction neutralization test; PRNT_50_, 50% plaque reduction as measured by probit regression. P value determined using Mann Whitney U test.

Neutralisation against MPXV Clade Ib and IIb was measured in non-heat-inactivated serum samples from the 25 vaccine recipients. The median PRNT_50_ value for Clade Ib was 25.9 (IQR = 10.05 – 49.7) compared to Clade IIb which had a median of 44.8 (IQR = 19.55 – 89.4). Comparisons across these samples demonstrated that two doses of MVA-BN generate greater neutralisation against MPXV Clade IIb than Ib, a difference which was found to be statistically significant with a p value of 0.0028 using a Wilcoxon signed-rank test (Figure 2).

**Figure 2.**
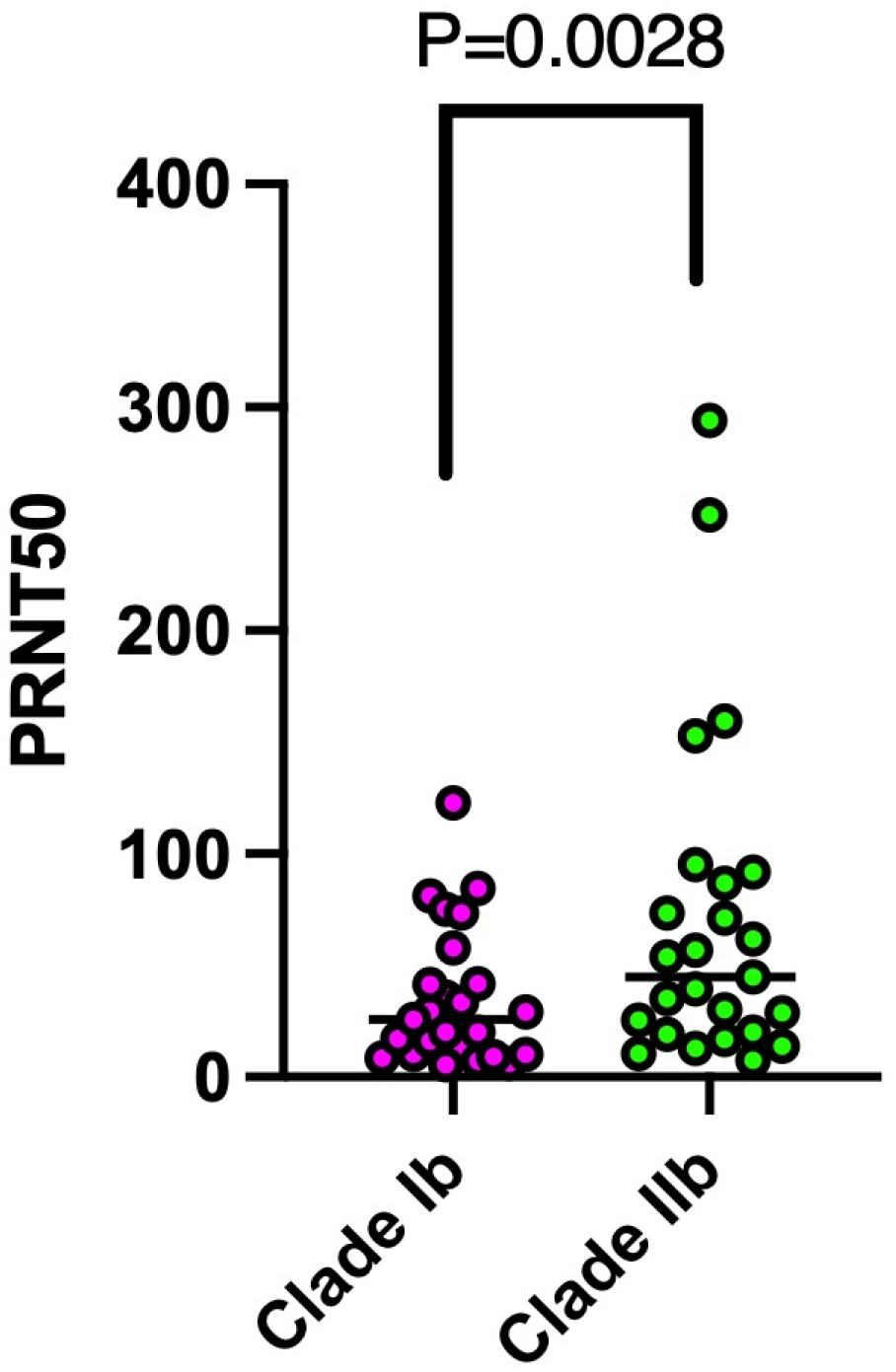
PRNT titres for participants vaccinated with 2 doses of MVA BN vaccine to assess neutralising antibody responses to MPXV Clade Ib and Clade IIb. Each datapoint represents the geometric mean titre of two experimental replicates. Black line represents median. MPXV, monkeypox virus; PRNT, plaque reduction neutralization test; PRNT_50_, 50% plaque reduction as measured by probit regression. P value determined using Wilcoxon matched pairs signed rank test.

## DISCUSSION

In this study we measured the levels of neutralising antibodies against MPXV Clades Ib and IIb in healthy adults vaccinated with two doses of MVA-BN. Our results demonstrate low levels of MPXV neutralisation; consistent with previous studies^3,4,5^. Neutralisation of Clade Ib was lower than Clade IIb. Although the present study is limited by the relatively low sample size it is the first to show neutralisation of MPXV Clade Ib in vaccine recipients who never had disease, and the first comparison with neutralisation of Clade IIb. Whether the low levels of neutralisation observed in this study correlates with protection is unclear, however in humans a single dose is known to confer protection in the short term and studies in animal models suggest MVA vaccination can provide cross protection against MPXV^10,11^. Moreover, given that the study cohort includes healthcare workers at highest risk of exposure evidence of vaccine-associated neutralisation is relevant for policies regarding future vaccine rollouts.

MPXV neutralisation is known to require complement^6^. Low levels of neutralisation were observed with the addition of guinea pig serum to virus without the addition of serum and when pooled human plasma was added to virus (data not shown) highlighting the non-specific effect that foreign complement sources can have on MPXV neutralisation. An effect which has also been observed with guinea pig serum alone exhibiting neutralization activity against mumps virus compared to purified antibodies^14.^ Therefore, like other studies^5^ an approach to use non-heat-inactivated serum to measure MPXV neutralisation was undertaken. The low levels of neutralisation identified in our study, particularly for Clade Ib, may suggest that at least moderate protection against disease due to MPXV Clade Ib from MVA-BN can be expected. The durability of these responses and whether a third ‘booster’ dose may be required to increase immunity to mpox Clade Ib infections^3,4,5^ requires further investigation.

## Data Availability

All data produced in the present study are available upon reasonable request to the authors.

## ACKNOWLEDGEMENTS

The authors would like to thank Dr Neil Blake, for his work on amplifying the Clade Ib strain.

This study was funded by The DECIPHER consortium consisting of The University of Liverpool, National Health Laboratory Services, Makerere University, Uganda National Health Research Organisation, McMaster University, Institut National De Recherche Biomedicale Du Zaire and Univesite Catholique De Bukavu. VS, CD and JH are based at the University of Liverpool.

This study was also funded by The Pandemic Institute, which is formed of seven founding partners: The University of Liverpool, Liverpool School of Tropical Medicine, Liverpool John Moores University, Liverpool City Council, Liverpool City Region Combined Authority, Liverpool University Hospital Foundation Trust, and Knowledge Quarter Liverpool. KS and LT are based at the University of Liverpool.

The International Severe Acute Respiratory and Emerging Infection Consortium (ISARIC) World Health Organization Clinical Characterisation Protocol (CCP-UK) United Kingdom is co-led and maintained by Prof. J Kenneth Baillie (Baillie Gifford Pandemic Science Hub, University of Edinburgh and Prof. Malcolm G Semple (Liverpool) on behalf of the ISARIC4C Investigators [isaric4c.net].

The views expressed are those of the author(s) and not necessarily those of the DECIPHER consortium, The Pandemic Institute or ISARIC4C Investigators.

