## Supplemental Table 1 for "Monkeypox virus-neutralising antibodies detected against Clade Ib and Clade IIb in healthy individuals following MVA-BN vaccination"

Supplemental Table 1. Demographics of the study cohort.

|  | **n** | **Age*** | **Ethnicity (%)** | **Sex (%)** | **Comorbidities** |
| --- | --- | --- | --- | --- | --- |
| **MVA-BN vaccine recipients** | 25 | 39  (30 – 45) | White (80%)  Asian (12%)  Latin (8%) | Male (36%)  Female (64%) | Multiple Sclerosis (n=1), Psoriasis (n=1), Asthma (n=1) |
