## Supplemental Figure 1 for "Monkeypox virus-neutralising antibodies detected against Clade Ib and Clade IIb in healthy individuals following MVA-BN vaccination"

Supplemental Figure 1. Phylogenetic tree of MPXV clades. Highlighted in red are the virus stock sequences mapped to corresponding clades.

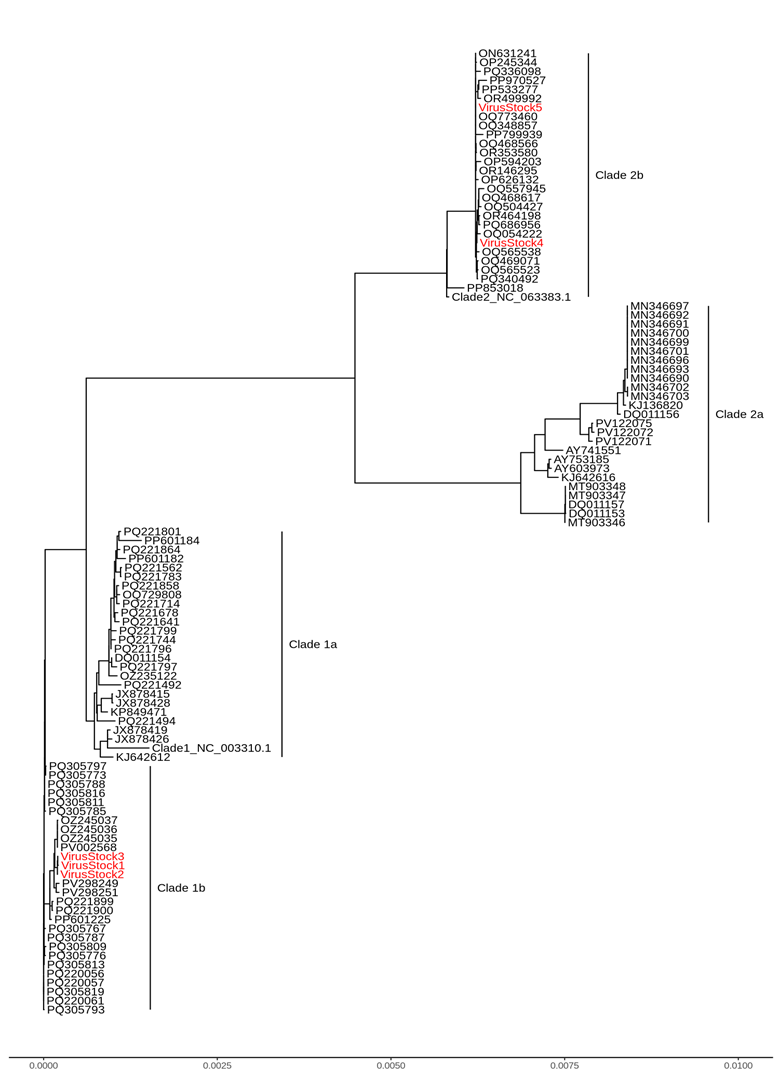
